# Deep learning segmentation model for automated detection of the opacity regions in the chest X-rays of the Covid-19 positive patients and the application for disease severity

**DOI:** 10.1101/2020.10.19.20215483

**Authors:** Haiming Tang, Nanfei Sun, Yi Li, Haoran Xia

## Abstract

**Purpose:** The pandemic of Covid-19 has caused tremendous losses to lives and economy in the entire world. The machine learning models have been applied to the radiological images of the Covid-19 positive patients for disease prediction and severity assessment. However, a segmentation model for detecting the opacity regions like haziness, ground-glass opacity and lung consolidation from the Covid-19 positive chest X-rays is still lacking.

**Methods:** The recently published collection of the radiological images for a rural population in United States had made the development of such a model a possibility, for the high quality images and consistent clinical measurements. We manually annotated 221 chest X-ray images with the lung fields and the opacity regions and trained a segmentation model for the opacity region using the Unet framework and the Resnet18 backbone. In addition, we applied the percentage of the opacity region over the area of the total lung fields for predicting the severity of patients.

**Results:** The model has a good performance regarding the overlap between the predicted and the manually labelled opacity regions. The performance is comparable for both the testing data set and the validation data set which comes from very diverse sources. However, careful manual examinations by experienced radiologists show mistakes in the predictions, which could be caused by the anatomical complexities. Nevertheless, the percentage of the opacity region can predict the severity of the patients well in regards to the ICU admissions and mortality.

**Conclusion:** In view of the above, our model is a successful first try in the development of a segmentation model for the opacity regions for the Covid-19 positive chest X-rays. However, additional work is needed before a robust model can be developed for the ultimate goal of the implementations in the clinical setting.

Model and supporting materials can be found in https://github.com/haimingt/opacity_segmentation_covid_chest_X_ray.

## 1 Introduction

Ever since the pandemic of Covid-19 in late 2019 till October 2020, there have been 38.8 million cases infected with this virus in the entire world, causing 1.1 million deaths. In the United States alone, 21.8 thousand patients had died of the infection and its complications [4]. The pandemic of Covid-19 has caused enormous losses to the lives and economy. Accurate and fast scanning of the lung situations for assessment of the disease progression has become a global-wise ubiquitous requirement. During the peak period, more than 70,000 new cases were diagnosed almost every day in the United States, 7-10% among them were hospitalized, and 2% of the diagnosed population have to be sent to emergent departments who needed further assessment for ICU admissions. The clinical environment in the era of Covid-19 pandemic has become very challenging with high-pressure and stress.

Clinicians and researchers have been devoting tremendous amounts of work for this unprecedented challenge. The field of diagnostic radiology has been a crucial area in clinical management of this disease. Clinicians have summarized the imaging characteristics of the Covid-19 infections for both the chest CT images and chest X rays[7], including ground glass opacities, consolidations, haziness and many others. Computer assisted segmentation and classification models are quickly studied[1] to evaluate their abilities to assist physicians and radiologists in assessing the radiological results.

Researchers have developed machine-learning models for Covid-19 related imaging studies. One category of these models is the classification model of lungs infected with the Covid-19 vs. those of other diseases using either the chest CT images or chest X-rays[18]. Some researchers have claimed very good results of using machine learning algorithms in differentiating Covid-19 radiological characteristics, and some even claimed better model results than junior radiologists, but just slightly worse than experienced radiologists[18]. There have also been projects on predicting the severity of the Covid-19 pneumonia[11]. For example, a research group manually curated a severity score for 96 chest X ray images[6], and used features from a model previously trained on a large data set of the non-Covid cases to develop a linear regression model to predict the severity of the opacities in the chest X-ray. Another research group came up with 6 severity scores for each chest X-ray image, separately for the upper, middle and lower regions of the left and right lung regions[15].

As far as we know about the most up to date publications, a segmentation model that can detect the location of the abnormalities for the Covid-19 positive chest X ray images is still lacking, while work has been done to successfully segment the lung regions[3, 10]. However, a recently published data set has made the development of such a segmentation model a possibility.

Recently, a research group made publicly available a data set of the 115 Covid-positive patients in the rural areas of the United States[8, 5]. The data set includes the chest X-ray images of patients, chest CT images, as well as the patient’s basic measurements and clinical prognosis like ICU admissions or mortality. In contrast to most of the machine-learning projects on Covid-19, which were developed from the images collected sparsely from the online resources or various publications, this data set is more consistent in image qualities and clinical data accuracy.

In this data set, the radiological data represents a population of patients who were diagnosed with Covid-19 in a specific region. Most of the images are chest X-rays and only a small portion of patients ever received a CT scan. Among these Chest X-ray images, as high as 30% have clear lungs without any abnormalities. Thus, this data set avoids the problem of over-representation of the more severe cases, which could be assembled from many different areas of the world. Using such a dataset helps develop a more robust model for a general population.

With the availability of this higher quality data, we aim to develop a model of segmentation for abnormal areas in chest X-ray images of the Covid-19 patients. More specifically, we want to locate the opacity regions that are characteristic of the Covid-19 infections, including haziness, ground glass opacities and consolidations. We want to answer two specific questions in this study: the first question is the development a model to point out the areas of the opacities for Covid-19 positive chest X-ray images, and other other question is the significance of the opacity info in determining the clinical severity of the patients.

## 2 Data and methods

### 2.1 Primary Data source

Recently, a collection of radiographic and CT images studies, together with patient demographics, comorbidities, prognosis and key radiology findings, as of July 2020, was published for a rural Covid-19 positive population in the southern United States[8]. The collection was downloaded from the Cancer Imaging Archive (TCIA) Public Access[5], containing 256 studies for a total number of 105 patients. Due to the focus of this study, we filtered out CT images and also X-rays that were taken in positions other than AP or PA. In total, we have included 221 chest X-ray images from 105 patients in our study. We randomly divided the entire dataset to the training and testing data sets by a ratio of 7:3 (154 images for the training set, 67 images for the testing set), but ensured that the images of the same patient were divided to the same subset.

### 2.2 Validation dataset

Cohen et al. published a chest X-ray dataset together with the severity scores[6]. The images in this collection were from various sources and serve as a good outside validation dataset. Due to the time consuming process of manual curations, we randomly selected a smaller dataset of 25 images from the original 96 images as our validation dataset.

### 2.3 Manual curations

Manual curations of the lung regions and the opacity regions were performed for all the images in the above training, testing and validation data sets. For simplicity considerations as well as the limitation of the resolution of the X-ray images, opacity regions were not differentiated among the ground glass opacities, haziness and consolidations. Lableme[17] was used for the curation work, and the opacity regions were represented by polygons of connected dots. The curations of the lung and opacity regions were saved in the Json format. Manual curations were performed by a junior physician and were reviewed by an experienced radiologist.

### 2.4 Image preprocessing and augmentation

The input X-ray image was then cropped to only keep the lung regions by mapping the original image with the lung contour segmentation. The model output was an image mask that has values 1 for manually curated opacity regions and 0 for all other regions.

Although the majority of our data comes from the same data source, the images were not of the same size. For easier deployment of our model, all training and testing images were resized to 320 by 320 pixels.

Data augmentation is a powerful technique to increase the amount of data and prevent model overfitting. Since our dataset is very small, we applied several different augmentations: horizontal flip, affine transforms, perspective transform, brightness/contrast/colors manipulations, image blurring and sharpening, gaussian noise and random crops. All of these transformations were performed using Albumentations, a fast augmentation library[2].

Sample images of the model input and output were included in Figure 1.

**Fig. 1.**
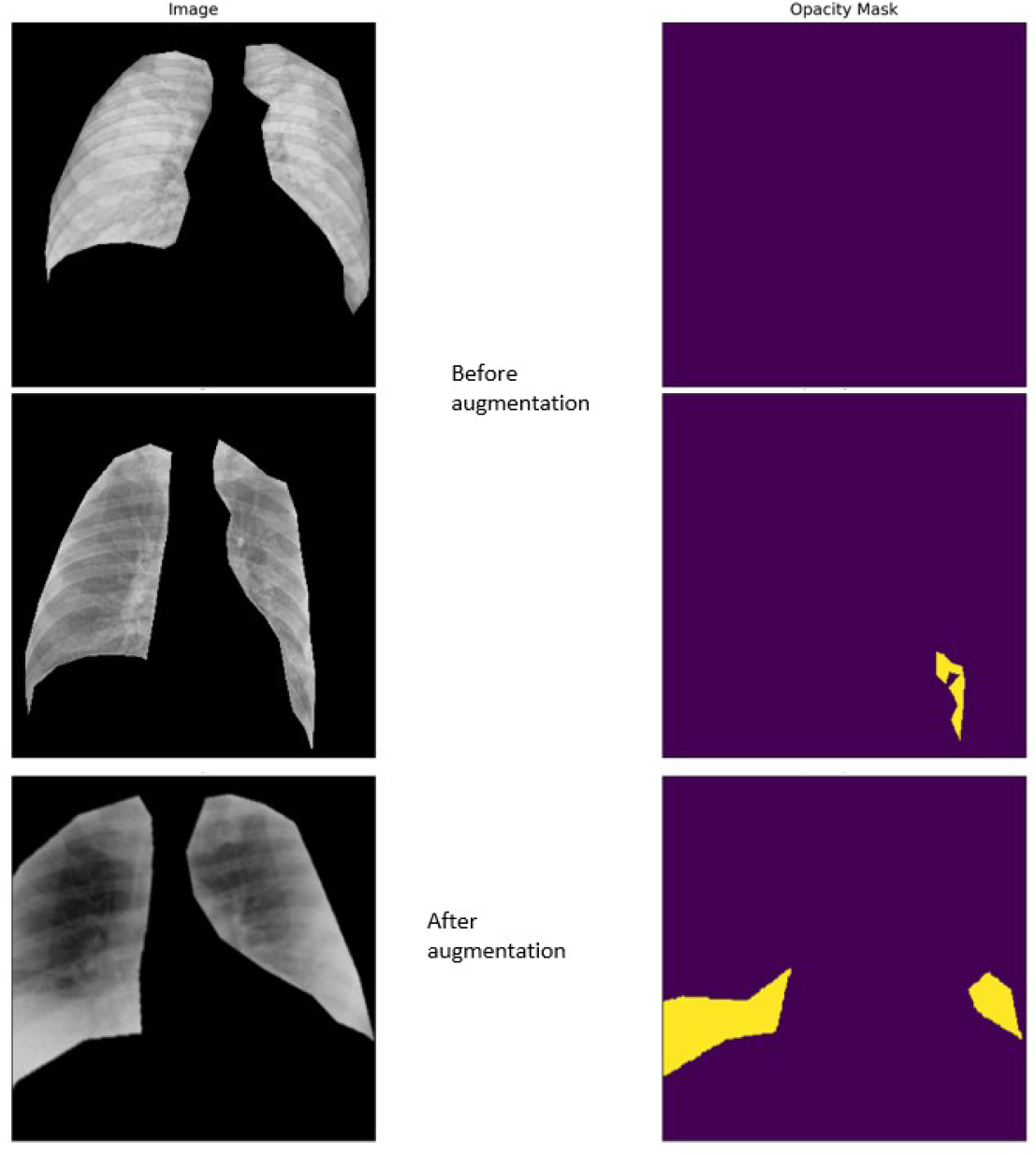
Sample images of the model input and output before and after augmentation.

### 2.5 Model architecture and training

“Segmentation models” is a python library with neural networks for image segmentation based on Keras and Tensorflow. It provides a high level API for segmentation model architectures and neural network backbones. For this study, we have chosen U-Net, the most popular and highly cited architecture which was developed specifically for biomedical image segmentation[14]. We have chosen the neural network backbone of ResNet18[9] for its wide usage, relatively smaller number of parameters and proven high performance. The topology of the adopted network was illustrated in the GitHub link. Segmentation loss was chosen to be Dice Loss[16] plus Focal Loss[12]. Dice Loss is essentially a measure of overlap between two samples, while Focal Loss is a measurement of how far off a prediction is from the truth.

### 2.6 Model metrics

We have chosen two popular metrics, IoU (Intersection over Union) and F1-score, to evaluate our model performance[13]. IoU is calculated using the area of overlap over the area of union between the input and prediction. F1-score is 2 * the Area of Overlap divided by the total number of pixels in both images. The model metrics were evaluated on several different datasets, including the training dataset alone, the testing dataset alone, the training and testing dataset combined (the whole dataset), the validation dataset, the images without opacity regions in the whole dataset as well as the images with opacity regions in the whole dataset.

### 2.7 Correlation with the disease severity

We calculated the percentage of opacity region by simply dividing the size of both lung fields by the size of the opacity regions. We then used this metric as the opacity percentage for predicting the severity of the patients, which was extracted from the excel table of the primary data source. The prognosis of each patient was among 3 categories: recovered with no ICU admission; recovered after ICU admission or deceased after ICU admission. For this data source, all patients who were deceased had gone to the ICU. The receiver operating curve (ROC) for the ICU admission was plotted using the opacity percentage against whether a patient was admitted to the ICU. The receiver operating curve (ROC) for the mortality was plotted using the opacity percentage against whether a patient was deceased.

We also downloaded a recently published model [6] for predicting the severity of the chest X-ray of the Covid-19 positive patients for a comparison. ROC curves for ICU admission as well as mortality were calculated in a similar way using the output opacity score of that model.

## 3 Results

### 3.1 Metrics of the trained model

The model metrics during the training process can be found in Figure 2. We observed a steady increase of the IoU score for the training dataset and a steady decrease of the model loss with increased epochs. The IoU score and the model loss for the testing dataset showed significant fluctuations during epochs 10 to 25, with a gradual return to a more steady state during epochs 30 to 50. In general, the trend of the metrics for both the training and the testing data sets was consistent, showing a gradual improvement in the training process.

**Fig. 2.**
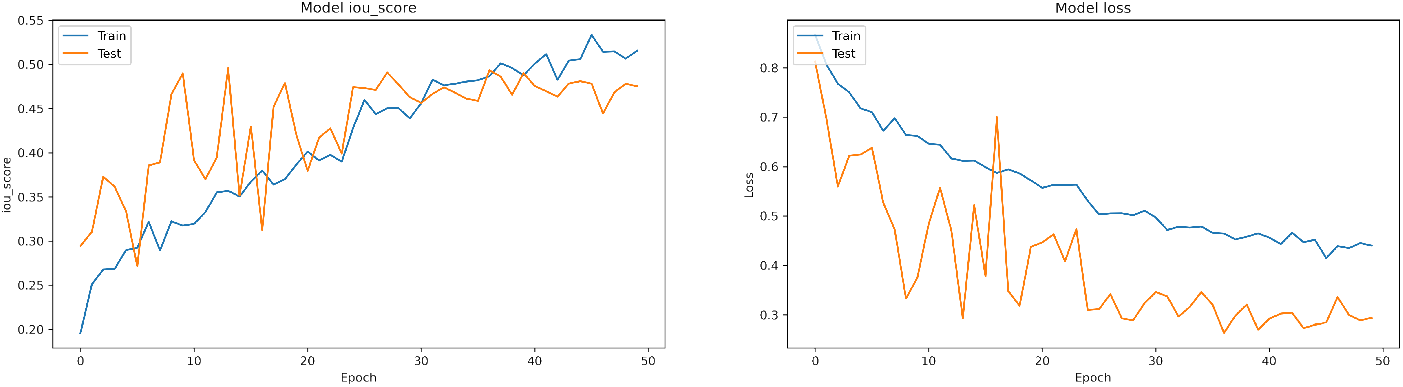
Model metric in the training process.

At the end of the training, the IoU score for the training dataset is 0.5157, while the IoU score for the testing dataset is 0.4755 and the IoU score the validation dataset is 0.6724. An IoU score >0.5 is normally considered a “good” prediction[13]. Although our measurement was very close to this threshold, thus, we had doubts about the actual performance of our model.

In the meantime, the validation dataset is from an alternative source. We usually expect the same or worse performance in the validation dataset as the model trained on a dataset with a limited variability may be lacking generalization capabilities to a dataset from an alternative source. The slightly more superior performance raises strong suspicions. These issues area covered in more detail in the discussion part.

### 3.2 Usage of the predicted opacities for predicting the disease severity

The ROCs of the opacity percentage on the mortality and the ICU admissions can be viewed in Figure 3.a and 3.b. We see a great predictability of the opacity percentage for the severity of the patients regarding mortality and the ICU admissions. This is consistent with our expectations, as the opacity region is the most intuitive measurement for assessing the patients by the physicians and radiologists in the clinical setting. In addition, multiple papers have quantitatively measured the associations of the opacity region with the disease severity in the clinical settings[18]. Thus, the results of our study reiterate this relationship. Besides, the consistency between the performances of our model predictions and the manually curated results show a reasonably well-performing model.

**Fig. 3.**
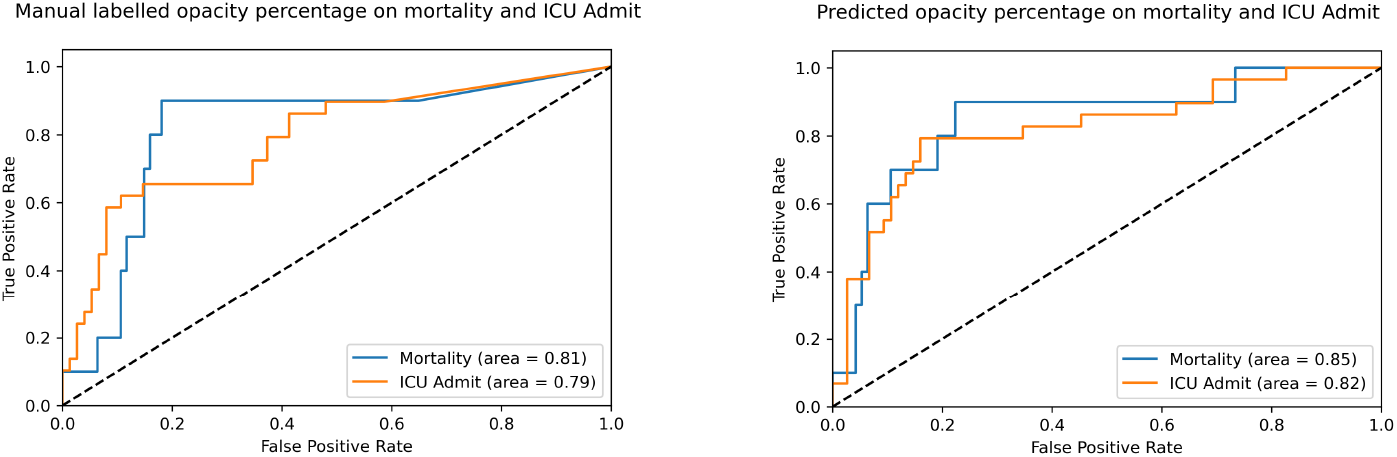
Performance of the opacity percentage in predicting mortality or ICU admission. 3.a, Opacity percentage calculated from manual labeling; 3.b, Opacity percentage calculated from model predictions.

### 3.3 Comparisons with the opacity score method

The Cohen group[6] developed a method to predict the severity of the chest X-ray of the Covid-19 infections. We ran this method on our data sets and applied the predicted opacity score for plotting the ROCs using the same patient severity data as above. The results can be viewed in Figure 4. We see a surprisingly good consistency between the performances of the opacity score of Cohen’s method and the opacity percentage of our model. The consistency is a confirmation of the good performance of our segmentation model. Considering the minimal simplicity of our metric, there could be a potential further development of a model for predicting the severity of the chest X-rays. However, this is not the scope of this study.

**Fig. 4.**
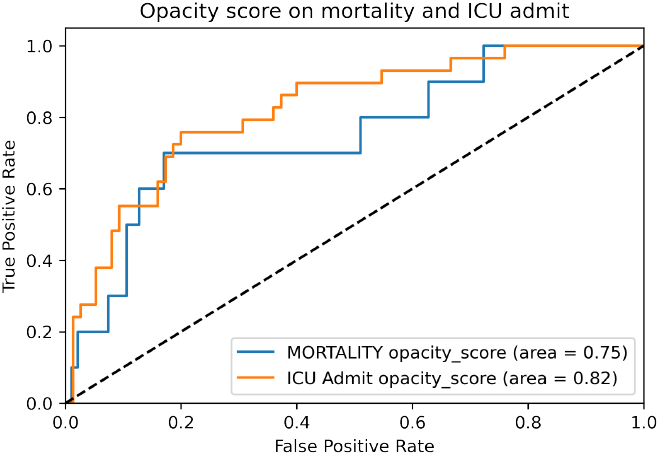
Performance of the opacity percentage in predicting mortality or ICU admission.

## 4 Discussions

### 4.1 The impact of the empty mask for IoU score

Due to the questions and suspicions we had for the IoU scores in the above, we further explored this issue and found a problem that significantly affected the IoU score. As defined by the formulas, the denominators are the union of the 2 regions for the IoU score. However, for our dataset, there are a significant number of images that do not have any opacity regions. These images have radiology reports of “clear lungs”, and the model predicted regions are also usually none. Thus, by definition, the union of the manually labelled region and the predicted region is 0, and a meaningful IoU score can not be calculated. To prove our hypothesis above, we extracted those images without any manually labelled opacity regions from the whole dataset combining the training and testing data sets. Then we re-applied the IoU score metrics to the new model created. It turned out that for the subset of 71 images without opacity regions, the calculated IoU score was 2.4E-10, close to 0. But for the rest 150 images with manually labelled opacity regions, the calculated IoU score increased to 0.6143 from the previous measurements of 0.5157 for the training dataset and 0.4755 for the testing set. Thus, the adjusted corrected IoU score for the entire dataset should be around (0.6143*150 + 1*71)/221 = 0.738, a much acceptable good performance for a segmentation model.

The superior performance of IoU score on the validation dataset, however, may come from the fact that most of the images in the validation dataset do contain manually labelled opacity regions. After the adjustment above, the IoU score of the validation dataset is slightly worse than that of the training and testing data sets.

This discrepancy may come from the differences in these dataset: our model was trained using the images from patients in the rural US regions, while the images of the validation data set are from sources that are significantly different.

### 4.2 Imperfections of the models

We printed out the predicted opacity regions for each of the images in our dataset for a manual examination. The comparison images are available in the supplemental materials. The examinations revealed some discrepancies, from which we found several potential shortcomings of this segmentation model.

Figure 5 lists several representative samples that have discrepancies between manual labeling and predictions.

**Fig. 5.**
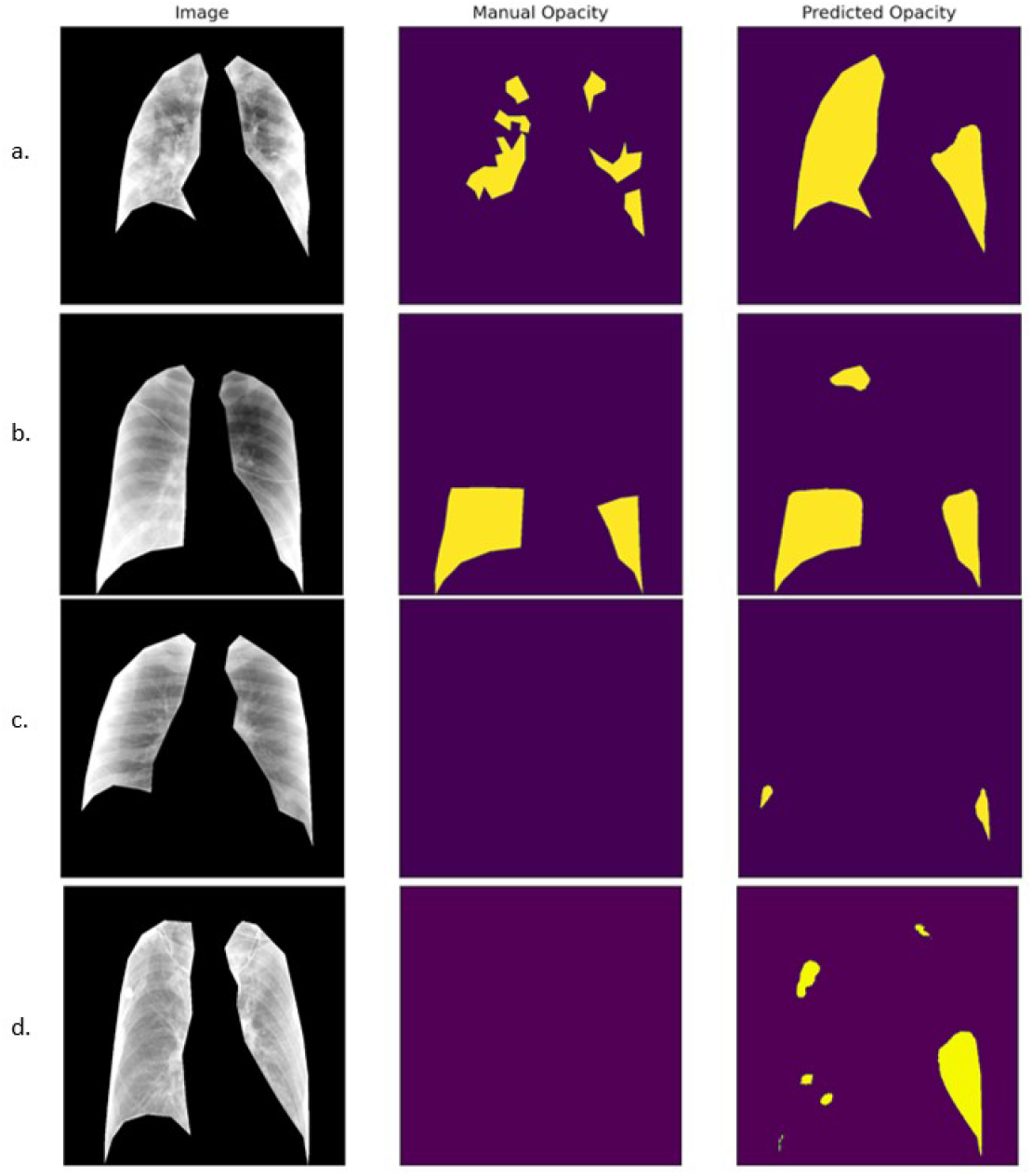
Representative samples that have discrepancies between manual labeling and predictions.

Firstly, the model flaw may come from the imperfections in the manual curations. Figure 5.a shows that the manual curated regions are very scattered, while the predicted regions cover larger and more consistent lung regions. This actually may represent the imperfections in the manual labeling process. This problem roots back to the ground truth for the opacity regions. This, however, is non-trivial. The boundary of the opacity region is very difficult to be clearly identified in the chest X-rays due to many issues, like image quality, patient anatomy, baseline disease etc. In the clinical setting, even experienced radiologists can identify a region of abnormality barely in a rough fashion. The clear delimitation of the abnormality boundaries are usually difficult, especially for lighter representations like ground glass opacity or haziness.

Secondly, some anatomical structures may compromise the model predictions. Figure 5.b shows that the chest X rays have opacities in both lung bases, however, the model prediction also contains a small region in the right lung apex. However, this is a wrong prediction probably caused by the rib shadow. Figure 5.c shows several tiny opacities in bilateral lung bases. However, the model may mistakenly predict the high density as an opacity. The high density is more likely due to the overlap of lung tissues instead of the ground glass opacities or consolidations caused by infections and exudates.

Thirdly, as shown in Figure 5.d, the model may mistake the markings of bronchial trees for opacities. The lungs in Figure 5.d are actually clear, but the markings of bronchial trees are prominent. They were likely to have been mistakenly predicted as opacities due to the higher densities.

## 5 Future improvements

As discussed above, the model predictions may be defective in a series of conditions. The segmentation of the opacity area in the chest X ray is a challenging work, even for experienced radiologists. The clear delimitation of the opacities is not as easy as delimitation of a solid mass with clear margins like tumors.

In general, the model performance is acceptable. Model metrics of IoU score and F1 score show good overlap between the manually labelled area and the predicted regions. From our examinations, correct locations can be detected more than 80% of the time, however, the exact boundaries may need further improvement.

To eliminate the influences of the anatomical structures or external instruments like ribs, scapula, wires and pacemakers, we probably need to include more chest X-ray images to more carefully label opacities. Another future direction is to develop a more robust model that takes into account all of these confounding elements. A possibility could be a multi-category segmentation model that detects the different anatomical structures separately.

## 6 Conclusion

In this study, we have demonstrated the development of a segmentation model for opacity regions for chest X-rays of the Covid-19 positive patients. The model performance is generally good with a precise predicted location for the opacities. Besides, the model shows a good generalization capacity when applied to a validation dataset composed of images of different sources. In addition, the percentage of the predicted opacity regions over the total lung area can predict the patient severity well regarding ICU admission and mortality. The performance of patient severity prediction is comparable or slightly better than the previously published “opacity score” method. In despite of these results, the model has a lot of imperfections in predicting the correct opacity regions. This may not only root from the lack of training data diversity, but also from the imperfections in manual labeling. Additional work is needed before a robust and accurate model can be developed for the ultimate goal of implementation in the clinical setting. In view of the above, our model is a successful first try in developing a segmentation model for the opacity regions for the Covid-19 positive chest X-rays. Our model schema and the manual segmentation data set may lay the foundation for the progress of more robust and accurate lung segmentation models in the future.

## Data Availability

The model, manual segmentation and other supporting materials can be found in https://github.com/haimingt/opacity_segmentation_covid_chest_X_ray.

## 7 Declarations

### 7.3 Ethics approval

Not applicable

### 7.4 Consent

Not applicable

### 7.5 Data and/or Code availability

https://github.com/haimingt/opacity segmentation covid chest X ray

### 7.6 Authors’ contribution statements

Haiming Tang (HT) and Nanfei Sun (NS) designed the study. HT performed manual segmentations of the chest X-ray images. Yi Li (YL) reviewed and corrected the segmentations. HT and Haoran Xia (HX) performed the model training, model validations and statistical analysis. HT and NS prepared the manuscripts. NS and YL reviewed the manuscripts.

## References

1. Abraham B, Nair MS (2020) Computer-aided detection of covid-19 from x-ray images using multi-cnn and bayesnet classifier. Biocybernetics and biomedical engineering 40(4):1436–1445, DOI 10.1016/j.bbe.2020.08.005

2. Buslaev A, Iglovikov VI, Khvedchenya E, Parinov A, Druzhinin M, Kalinin AA (2020) Albumentations: Fast and flexible image augmentations. Information 11(2), DOI 10.3390/info11020125

3. Candemir S, Antani S (2019) A review on lung boundary detection in chest X-rays. International Journal of Computer Assisted Radiology and Surgery 14(4):563–576, DOI 10.1007/s11548-019-01917-1

4. Centers for Disease Control and Prevention (2020 (accessed October 16, 2020)) Covidview: A weekly surveillance summary of u.s. covid-19 activity. URL https://www.cdc.gov/coronavirus/2019-ncov/covid-data/covidview/index.html

5. Clark K, Vendt B, Smith K, Freymann J, Kirby J, Koppel P, Moore S, Phillips S, Maffitt D, Pringle M, Tarbox L, Prior F (2013) The Cancer Imaging Archive (TCIA): maintaining and operating a public information repository. Journal of digital imaging 26(6):1045–1057, DOI 10.1007/s10278-013-9622-7

6. Cohen JP, Dao L, Morrison P, Roth K, Bengio Y, Shen B, Abbasi A, Hoshmand-Kochi M, Ghassemi M, Li H, Duong TQ (2020) Predicting covid-19 pneumonia severity on chest x-ray with deep learning. 2005. 11856

7. Cozzi D, Albanesi M, Cavigli E, Moroni C, Bindi A, Luvará S, Lucarini S, Busoni S, Mazzoni LN, Miele V (2020) Chest X-ray in new Coronavirus Disease 2019 (COVID-19) infection: findings and correlation with clinical outcome. La Radiologia medica 125(8):730–737, DOI 10.1007/s11547-020-01232-9

8. Desai S, Baghal A, Wongsurawat T, Al-Shukri S, Gates K, Farmer P, Rutherford M, Blake G, Nolan T, Powell T, Sexton K, Bennett W, Prior F (2020) Data from chest imaging with clinical and genomic correlates representing a rural covid-19 positive population. The Cancer Imaging Archive (TCIA) URL https://doi.org/10.7937/tcia.2020.py71-5978

9. He K, Zhang X, Ren S, Sun J (2016) Deep residual learning for image recognition. In: 2016 IEEE Conference on Computer Vision and Pattern Recognition (CVPR), pp 770–778, DOI 10.1109/CVPR.2016.90

10. Kholiavchenko M, Sirazitdinov I, Kubrak K, Badrutdinova R, Kuleev R, Yuan Y, Vrtovec T, Ibragimov B (2020) Contour-aware multi-label chest X-ray organ segmentation. International Journal of Computer Assisted Radiology and Surgery 15(3):425–436, DOI 10.1007/s11548-019-02115-9

11. Li MD, Arun NT, Gidwani M, Chang K, Deng F, Little BP, Mendoza DP, Lang M, Lee SI, O’Shea A, Parakh A, Singh P, Kalpathy-Cramer J (2020) Automated Assessment and Tracking of COVID-19 Pulmonary Disease Severity on Chest Radiographs using Convolutional Siamese Neural Networks. Radiology: Artificial Intelligence 2(4):e200.079, DOI 10.1148/ryai.2020200079

12. Lin T, Goyal P, Girshick R, He K, Dollár P (2017) Focal loss for dense object detection. In: 2017 IEEE International Conference on Computer Vision (ICCV), pp 2999–3007, DOI 10.1109/ICCV.2017.324

13. Rahman MA, Wang Y (2016) Optimizing intersection-over-union in deep neural networks for image segmentation. In: Advances in Visual Computing, Springer International Publishing, Cham, pp 234–244

14. Ronneberger O, Fischer P, Brox T (2015) U-net: Convolutional networks for biomedical image segmentation. In: Medical Image Computing and Computer-Assisted Intervention – MICCAI 2015, Springer International Publishing, Cham, pp 234–241

15. Signoroni A, Savardi M, Benini S, Adami N, Leonardi R, Gibellini P, Vaccher F, Ravanelli M, Borghesi A, Maroldi R, Farina D (2020) End-to-end learning for semiquantitative rating of covid-19 severity on chest x-rays. 2006.04603

16. Sudre C, Li W, Vercauteren T, Ourselin S, Cardoso MJ (2017) Generalised dice overlap as a deep learning loss function for highly unbalanced segmentations. DOI 10.1007/978-3-319-67558-928

17. Wada K (2016) labelme: Image Polygonal Annotation with Python. URL https://github.com/wkentaro/labelme

18. Zhang K, Liu X, Shen J, Li Z, Sang Y, Wu X, Zha Y, Liang W, Wang C, Wang K, Ye L, Gao M, Zhou Z, Li L, Wang J, Yang Z, Cai H, Xu J, Yang L, Cai W, et al (2020) Clinically applicable ai system for accurate diagnosis, quantitative measurements, and prognosis of covid-19 pneumonia using computed tomography. Cell 181(6):1423 – 1433.e11, DOI https://doi.org/10.1016/j.cell.2020.04.045

